# Estimation of COVID-19 recovery and decease periods in Canada using machine learning algorithms

**DOI:** 10.1101/2021.07.16.21260675

**Authors:** Subhendu Paul, Emmanuel Lorin

## Abstract

We derive a novel model escorted by large scale compartments, based on a set of coupled delay differential equations with extensive delays, in order to estimate the incubation, recovery and decease periods of COVID-19, and more generally any infectious disease. This is possible thanks to machine learning algorithms applied to publicly available database of confirmed corona cases, recovered cases and death toll. In this purpose, we separate i) the total cases into 14 groups corresponding to 14 incubation periods, ii) the recovered cases into 406 groups corresponding to a combination of incubation and recovery periods, and iii) the death toll into 406 groups corresponding to a combination of incubation and decease periods. In this paper, we focus on recovery and decease periods and their correlation with the incubation period. The estimated mean recovery period we obtain is 22.14 days (95% Confidence Interval(CI): 22.00 to 22.27), and the 90th percentile is 28.91 days (95% CI: 28.71 to 29.13), which is in agreement with statistical supported studies. The bimodal gamma distribution reveals that there are two groups of recovered individuals with a short recovery period, mean 21.02 days (95% CI: 20.92 to 21.12), and a long recovery period, mean 38.88 days (95% CI 38.61 to 39.15). Our study shows that the characteristic of the decease period and the recovery period are alike. From the bivariate analysis, we observe a high probability domain for recovered individuals with respect to incubation and recovery periods. A similar domain is obtained for deaths analyzing bivariate distribution of incubation and decease periods.

## Main

The outbreak of coronavirus disease 2019 (COVID-19), reported early in Wuhan (China)^1^ and spread around the world, is creating dramatic and daily changes with profound impacts worldwide. As a consequence the outbreak was declared a pandemic by the World Health Organization (WHO) in March 2020^2^, and by the end of 2020, COVID-19 has infected about 79.2 millions of people in the world, with an approximate cumulative global mortality of 3.2%^2^. To limit the impact of this deadly virus, a rapid and widespread vaccination of the population is now in place. However, it is established that vaccine are not 100% effective to stop the transmission or infection of COVID-19. In addition, huge numbers of global SARS-CoV-2 infections have led to the emergence of variants, notably Alpha (B.1.1.7 UK), Beta (B.1.351 S. Africa), Gamma (P.1 Brazil), Epsilon (B.1.429 California), Iota (B.1.526 New York), Delta and Kappa (B.1.617.2 and B.1.617.1 India) which make the situation more challenging. In this circumstance to get a complete feature of COVID-19, it is essential to fully understand the key (incubation, recovery and decease) periods.

We already successfully estimated the incubation period of COVID-19 in Canada^3^. In the present context, we focus on the recovery and decease periods and their correlation with the incubation period. In the current framework, we define the recovery period as the time from the contraction of the coronavirus to recovery, i.e., the incubation period plus the onset time from the symptom to recovery; the latter is the same as the viral shedding of SARS-CoV-2. We describe the decease period in the same way as the recovery period. Understanding the recovery period of disease is very useful information in the struggle against the disease. If the incidence of a disease is remarkably high and the recovery period of the disease is also high then the prevalence of the disease in the country is likely to increase which in turn puts extra health, economic and social burden on this country. Understanding the recovery period of the disease will help governments to plan proper strategies to counter the disease and to organize the requirements such as hospitals, doctors, medical staffs, medical equipment’s, etc. It will also help to implement different social and economic policies which will be essential to fight the disease.

There are several statistical studies^4–11^, based on various samples of patients such as severe, non-severe, ICU, non-ICU, large size, small size, meta-analysis, estimated the recovery time of the current pandemic. In addition to those statistical approaches, there are numerous analytical and computational studies based on mathematical models, involving Ordinary Differential Equations (ODE)^12–20^ as well as Delay Differential Equations (DDE)^21–26^, to calculate the basic reproduction number *R*_0_ and understand the underlying dynamics of the epidemic. Researchers usually consider single-delay models, occasionally two delays.

To the best of our knowledge, we demonstrate for the first time a substantial compartment based model, with a total 830 partitions, in order to estimate the key (incubation, recovery, decease periods) periods of COVID-19 as well as the bivariate distribution of incubation and recovery periods, and the bivariate distribution of incubation and decease periods. This will be achieved using publicly available database^27^ of the total number of corona-positive cases, recovery and death toll. Using the novel model, demonstrated here, we divide the publicly available database into thousands of groups, and these separated classes are the key source for estimating all the key periods. This approach is free from any special type of samples in order to produce the distributions of those periods; it only involves large scale computations for estimating about thousand model parameters. After a single calculation of this method, we can generate the current distributions as well as previous distributions of those periods. In the statistical based approaches, it is usually difficult to consider large incubation, recovery and decease periods if the sample size is small. However, in our approach, we can go well beyond 14 days, the maximum incubation period that we have set in this paper, and beyond the interval 2 weeks to 6 weeks, the range of recovery as well as decease periods that we have considered in the current computations. As of May 23, 2021, the World Health Organization (WHO) had confirmed a total of 1,359,180 cases of COVID-19 in Canada, including 25,231 deaths^2^. As of May 23, 2021 there are five provinces in Canada with death toll more than 1000 (Fig.1(a)), and the recovery and death rates are respectively 96.1% and 0.7% (Fig.1(c)). During the first wave of COVID-19 in Canada, January 22, 2020 to July 16, 2020, the recovery and death rates were respectively 66.5% and 8.1% (Fig. 1(b)). Here, we assume that the recovery and decease periods of COVID-19 remain unchanged i.e., these periods during the first wave and the present time are almost identical, and under this assumption we merely consider the database of first wave for the calculation.

**Figure 1.**
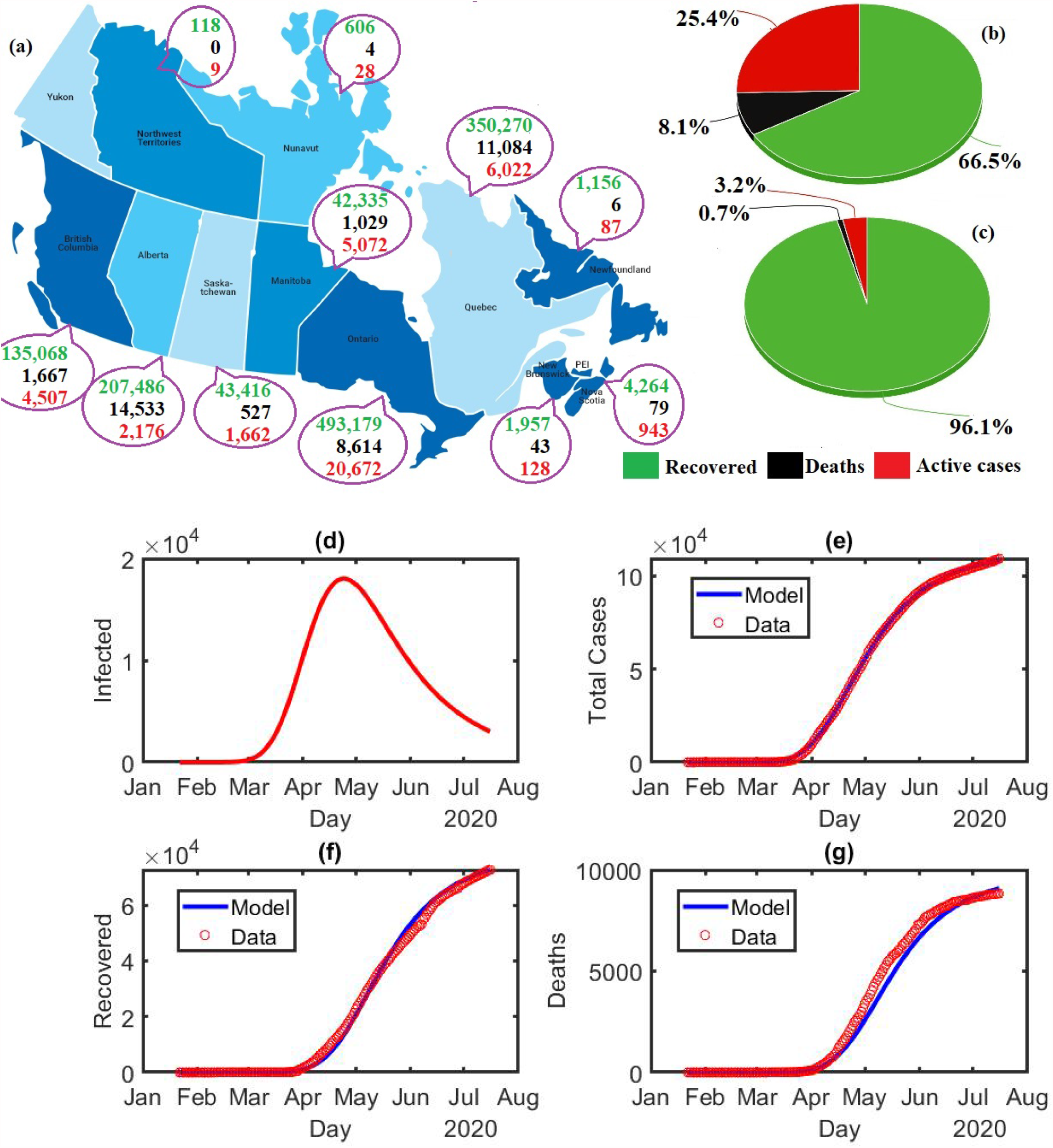
COVID-19 pandemic in Canada: **(a)** As of May 23, 2021 Canadian out break at-a-glance; the green, black and red digits are represented the number of recovered individuals, death toll and active cases, respectively. **(b)** Percentage of recovered (green), deaths (black) and active cases (red) in Canada during the first wave, January 22, 2020 to July 16, 2020. **(c)** Percentage of recovered (green), deaths (black) and active cases (red) in Canada as of May 23, 2021. **Model calculation for Canada during the first wave, January 22, 2020 to July 16, 2020: (d)** Estimation of the number of infected individuals. **(e)** Estimation of the total number of coronavirus cases compared to the available data^27^. **(f)** Estimation of the total number of recovered compared to the available data^27^. **(g)** Estimation of the total number of deaths compared to the available data^27^.

There are various studies on recovery period, and no result is reported (to the best of our knowledge) on bivariate distributions as mentioned above. The key periods may depend on age^28^ (median-age */* country), hard immunity, public health system, corona testing capacities, daily corona cases, etc. For a better estimation of the key periods for a particular region, we need to study local patients. Data collection is a bottleneck in studying those key periods for COVID-19 or other infectious diseases using clinical survey, and we need a sample of large size for bivariate analysis. However, key periods can easily be estimated using the approach we propose here, the publicly available database along with machine learning algorithms.

## Results

The proposed model assists us to generate new refined recovery and death toll database, *R*_*ik*_ and *D*_*ik*_, by dividing the total recovered individuals and the total number of deaths as of July 16, 2020 into myriad of groups. The new database is the key source for studying all kinds of distributions, reported in the article.

### Validation of the proposed model

After estimating the model parameters with sufficiently small values of error functions, we obtain a good agreement (Figs.1(e), 1(f) and 1(g)) between the calculated values of the model variables such as total corona-positive cases, number of recovered individuals, etc. and the available data^27^. The population of the infected group gradually increased until end of April 2020, and thereafter slowed down (Fig. 1(d)).

### Univariate distributions

The groups of recovered individuals *R*_*ik*_, *i* = 1, 2, …, 14 and *k* = 1, 2, …, 29, corresponding to the incubation period (in days) *τ*_*i*_, 1≤ *τ*_*i*_ ≤14, and recovery period (in days) *ζ*_*k*_, 14 ≤*ζ*_*k*_ ≤42 can be represented in a matrix form (Fig. 2(a)). We use the data set 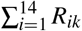 for *ζ*_*k*_ = 14, 15, …, 42 to obtain the frequency distribution for recovery period and the corresponding fitted gamma distributions, unimodal (Fig.2(b)) Γ(*ζ, K*_*r*_, *θ*_*r*_) and bimodal (Fig.2(c)) 0.9365 Γ(*ζ, K*_*r*1_, *θ*_*r*1_) + 0.0635 Γ(*ζ, K*_*r*2_, *θ*_*r*2_). Here, the variable *ζ* indicates the recovery period and the parameters *K*_*r*_ = 18.62067, *θ*_*r*_ = 1.18892, *K*_*r*1_ = 34.55447, *θ*_*r*1_ = 0.60847, *K*_*r*2_ = 226.40545 and *θ*_*r*2_ = 0.17171 with statistical *p* value less than 0.01. The mean recovery period we obtain using an unimodal gamma distribution is 22.14 days (95% CI 22.00 to 22.27); the median of the recovery period is 21.74 days (95% CI 21.61 to 21.87); the 90th percentile is 28.91 days (95% CI 28.71 to 29.13); the 95th percentile is 31.20 days (95% CI 30.95 to 31.45). For a better estimation, we use a bimodal distribution, a linear combination of Γ(*ζ, K*_*r*1_, *θ*_*r*1_) and Γ(*ζ, K*_*r*2_, *θ*_*r*2_). The mean of Γ(*ζ, K*_*r*1_, *θ*_*r*1_) and Γ(*ζ, K*_*r*2_, *θ*_*r*2_) are 21.02 days (95% CI 20.92 to 21.12) and 38.88 days (95% CI 38.61 to 39.15), respectively. The percentile curves of unimodal and bimodal gamma distributions show (Fig.2(d)) that the median of unimodal and bimodal are the same, although there are slight differences other than the median.

**Figure 2.**
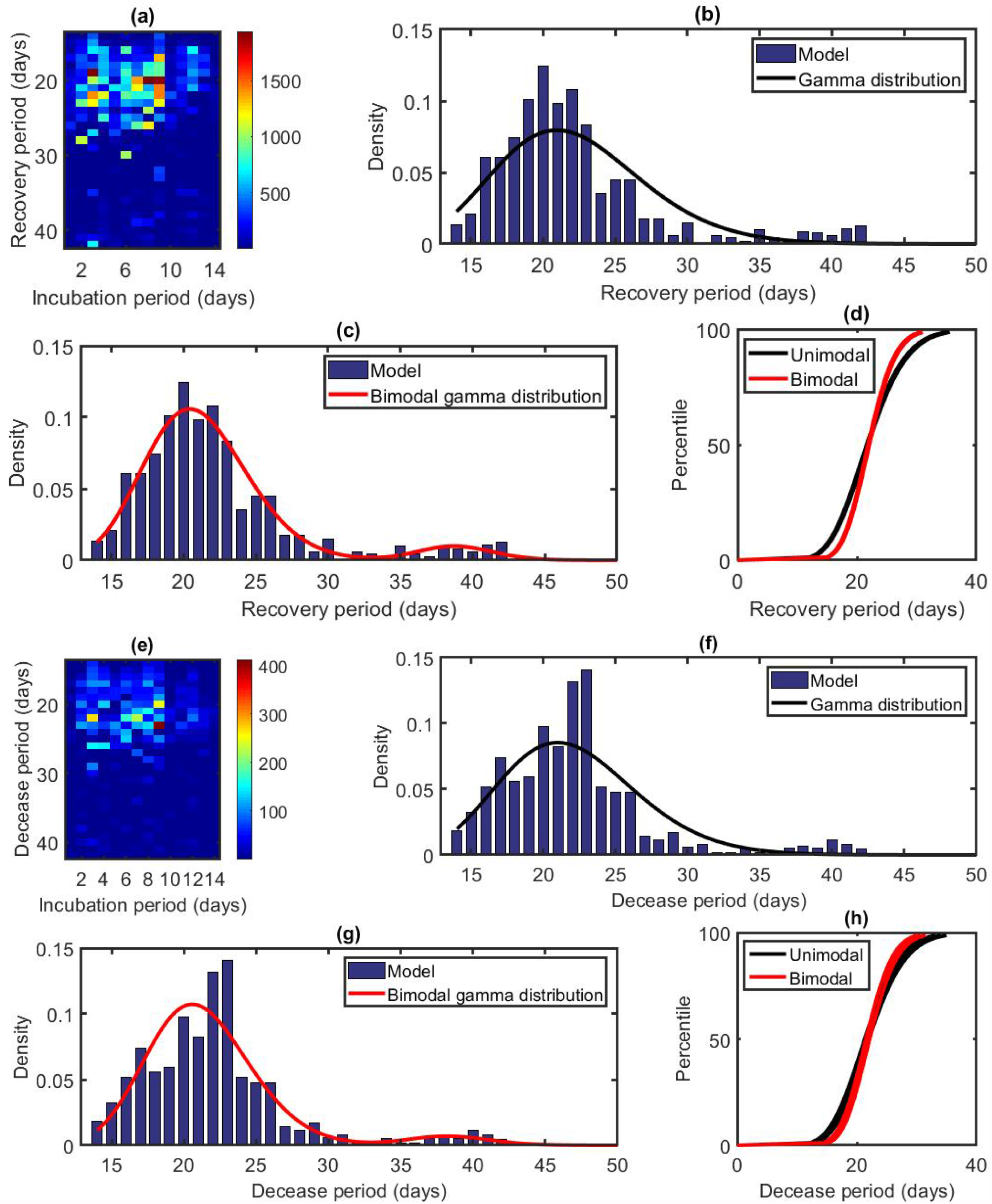
Distribution of the recovery and decease periods: Results based on the total recovered cases of the first 177 days during the pandemic in Canada starting from January 22, 2020 i.e., cumulative data as of July 16,2020. **(a)** Splitting values of recovered individuals as a function of incubation and recovery periods. **(b)** Probability density function of the gamma distribution *Γ(ζ, K, θ*) with *K* = 18.62067 and *θ* = 1.18892. The blue bars indicate the densities obtained from the model calculation. **(c)** Probability density function of the bimodal gamma distribution 0.9365 Γ(*ζ, K*_1_, *θ*_1_) + 0.0635 Γ(*ζ, K*_2_, *θ*_2_) with *K*_1_ = 34.55447, *θ*_1_ = 0.60847, *K*_2_ = 226.40545 and *θ*_2_ = 0.17171. The blue bars indicate the densities obtained from the model calculation. **(d)** Percentile curves for unimodal and bimodal gamma distributions. **(e)** Splitting values of the deaths as a function of incubation and decease periods. **(f)** Probability density function of the gamma distribution Γ(*η, K, θ*) with *K* = 21.33660 and *θ* = 1.03174. The blue bars indicate the densities obtained from the model calculation. **(g)** Probability density function of the bimodal gamma distribution 0.9508 Γ(*η, K*_1_, *θ*_1_) + 0.0492 Γ(*η, K*_2_, *θ*_2_) with *K*_1_ = 35.00855, *θ*_1_ = 0.60511, *K*_2_ = 186.11379 and *θ*_2_ = 0.20636. The blue bars indicate the densities obtained from the model calculation. **(h)** Percentile curves for unimodal and bimodal gamma distributions.

The groups death toll groups *D*_*ik*_, *i* = 1, 2, …, 14 and *k* = 1, 2, …, 29, and corresponding incubation period (in days)*τ*_*i*_, 1 ≤ *τ*_*i*_ ≤ 14, and decease period (in days) *η*_*k*_, 14 ≤ *η*_*k*_ ≤ 42 can be represented as a matrix (Fig. 2(e)). We use the data set 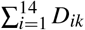 for *η*_*k*_ = 14, 15, …, 42 to obtain the frequency distribution for decease period and corresponding fitted gamma distributions, unimodal (Fig.2(f)) Γ(*η, K*_*d*_, *θ*_*d*_) and bimodal (Fig.2(g)) 0.9508 Γ(*η, K*_*d*1_, *θ*_*d*1_) + 0.0492 Γ(*η, K*_*d*2_, *θ*_*d*2_). Here, the variable *η* indicates the decease period and the parameters *K*_*d*_ = 21.33660, *θ*_*d*_ = 1.03174, *K*_*d*1_ = 35.00855, *θ*_*d*1_ = 0.60511, *K*_*d*2_ = 186.11379 and *θ*_*d*2_ = 0.20636 with statistical *p* value less than 0.01 for Γ(*η, K*_*d*_, *θ*_*d*_), Γ(*η, K*_*d*1_, *θ*_*d*1_) and equal to 0.18 for Γ(*η, K*_*d*2_, *θ*_*d*2_). The mean decease period we obtain using an unimodal gamma distribution is 22.01 days (95% CI 21.64 to 22.39); the median of the decease period is 21.67 days (95% CI 21.31 to 22.04); the 90th percentile is 28.30 days (95% CI 27.72 to 28.89); the 95th percentile is 30.39 days (95% CI 29.71 to 31.10). For better estimation, we use a bimodal distribution, a linear combination of Γ(*η, K*_*d*1_, *θ*_*d*1_) and Γ(*η, K*_*d*2_, *θ*_*d*2_). The mean of Γ(*η, K*_*d*1_, *θ*_*d*1_) and Γ(*η, K*_*d*2_, *θ*_*d*2_) are 21.18 days (95% CI 20.90 to 21.47) and 38.41 days (95% CI 37.41 to 39.40), respectively. The percentile curves show (Fig.2(h)) that the percentiles of unimodal and bimodal distributions are almost the same.

### Bivariate distributions

To analyze the bivariate distribution, we use the software Statgraphics^29^, based on the statistical package **R**. Using the elements *R*_*ik*_ for *i* = 1, 2, …, 14 and *k* = 1, 2, …, 29, we obtain a bivariate histogram (Fig.3(a)) for the incubation and recovery periods. There are two peaks at the points (3, 19), i.e., for *τ*_*i*_ = 3 and *ζ*_*k*_ = 19, and (8, 20), i.e., for *τ*_*i*_ = 8 and *ζ*_*k*_ = 20, corresponding to the high densities of recovered individuals. We estimate the histogram using a bivariate normal distribution 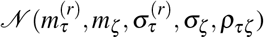 (Fig.3(b)) where the variables *τ* and *ζ* represent the incubation and recovery periods, respectively. The mean 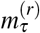 and standard deviation 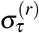 of the incubation period are 6.43 (95% CI 6.27 to 6.59) and 3.06 (95% CI 2.96 to 3.18), respectively; the mean *m*_*ζ*_ and standard deviation *σ*_*ζ*_ of the recovery period are 21.91 (95% CI 21.63 to 22.18) and 5.33 (95% CI 5.14 to 5.53), respectively; the correlation between incubation and recovery periods *ρ*_*τζ*_ is -0.11. The two dimensional representation of the bivariate normal distribution (Fig.3(c))shows that the highly probable recovery region (red in the figure) is a nested domain of *τ* = 6.43 and *ζ* = 21.91. To precisely analyze the highly probable region, we estimate the histogram (Fig.3(a)) using a nonparametric density function with a width of 33%, low and high percentage give a more local and global estimation, respectively, and we obtain a distribution with two peaks (Fig.3(d)). Two distinguishable peaks indicate that there are two separate highly probable regions surrounding the points *τ* = 3.49, *ζ* = 20.52 and *τ* = 8.38, *ζ* = 20.35 (Fig.3(e)). The bivariate mixture distribution analysis shows that we can estimate the histogram of *R*_*ik*_ for *i* = 1, 2, …, 14 and *k* = 1, 2, …, 29 using a combination of two bivariate normal distributions, 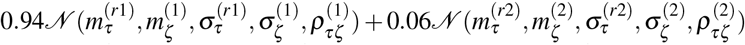 where the superscript 1 (resp. 2) represents the parameters for the first (resp. second) component, respectively. The parameters of the first component are 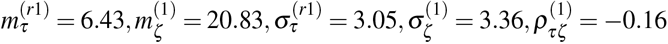, and those of the second component are 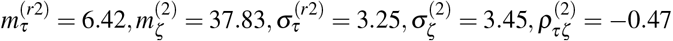.

**Figure 3.**
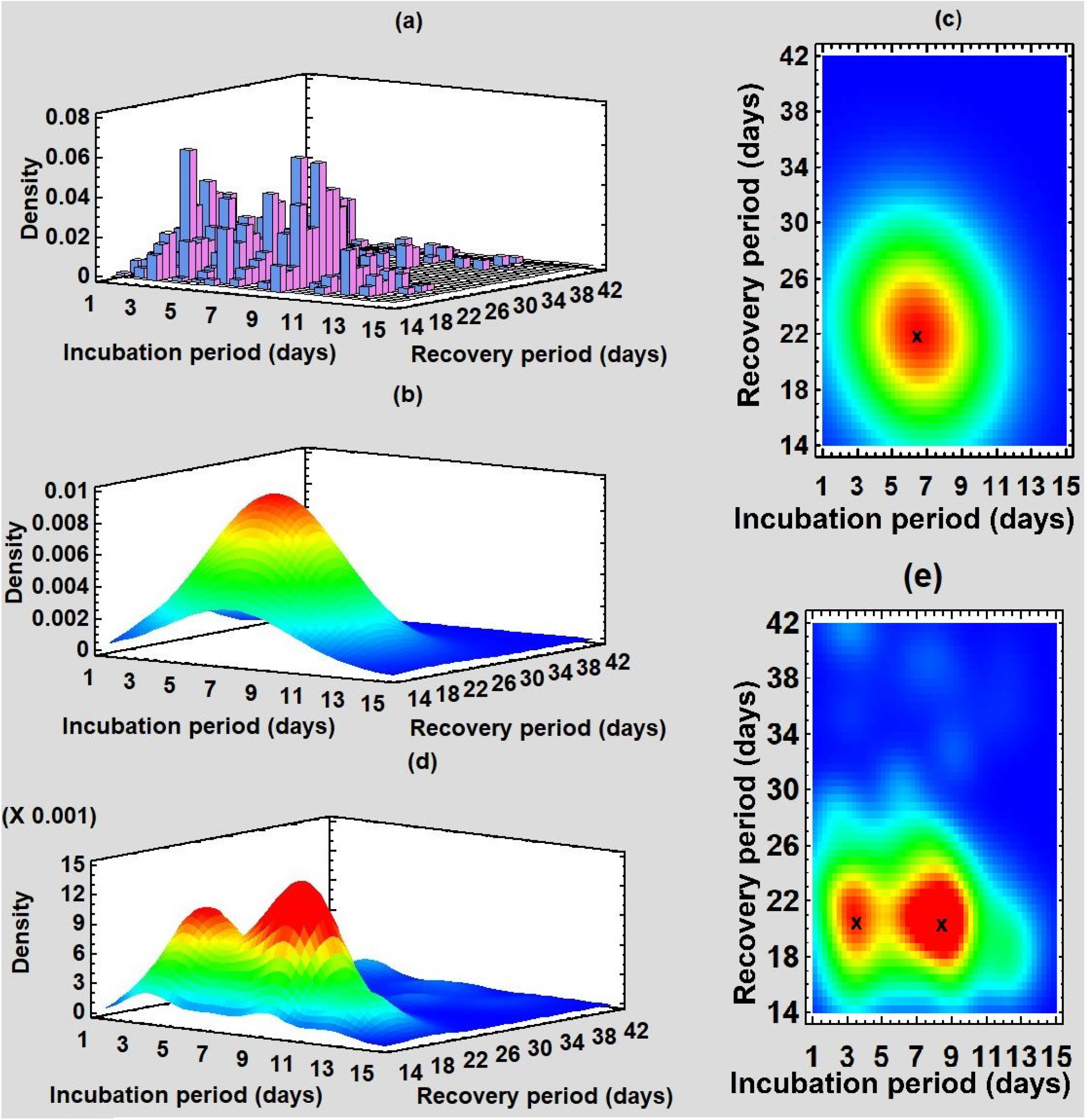
Bivariate distribution of the incubation and recovery periods: **(a)** Histogram of the estimated data *R*_*ik*_ for *i* = 1, 2, …, 14 and *k* = 1, 2, …, 29 using the model. **(b)** Fitted bivariate normal distribution. **(c)** Two-dimensional display of (b); the red region is the highly probable domain for recovery, and x (6.43, 21.91) denotes the center of the region. **(d)** Fitted nonparametric density estimate with wide 33%; two peaks show that there are two distinguishable high probable regions. **(e)** Two-dimensional display of (d); two x represent the centers of two high probable regions (3.49, 20.52) and (8.38, 20.35).

Using the elements *D*_*ik*_ for *i* = 1, 2, …, 14 and *k* = 1, 2, …, 29, we obtain a bivariate histogram (Fig.4(a)) for the incubation and decease periods. There are two peaks at the points (3, 22), i.e., for *τ*_*i*_ = 3 and *η*_*k*_ = 22, and (9, 23), i.e., for *τ*_*i*_ = 9 and *η*_*k*_ = 23, corresponding to the high densities of deaths. We estimate the histogram using a bivariate normal distribution 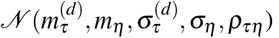 (Fig.4(b)) where the variables *τ* and *η* represent the incubation and decease periods, respectively. The mean 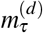 and standard deviation 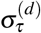 of the incubation period are 6.56 (95% CI 6.36 to 6.76) and 3.00 (95% CI 2.86 to 3.15), respectively; the mean *m*_*η*_ and standard deviation *σ*_*η*_ of the decease period are 21.64 (95% CI 21.33 to 21.94) and 4.43 (95% CI 4.23 to 4.65), respectively; the correlation between incubation and decease periods *ρ*_*τη*_ is -0.008. The two dimensional representation of the bivariate normal distribution (Fig.4(c)) shows that the highly probable decease region (red in the figure) is a nested domain of *τ* = 6.56 and *η* = 21.64. To precisely analyze the highly probable regions, we estimate the histogram (Fig.4(a)) using a nonparametric density function with a width of 40%, low and high percentage give a more local and global estimation, respectively, and obtain a distribution with two peaks (Fig.4(d)), one in the high probability region (red in figure) and another one in the second high probability region (yellow in figure). The highly probable region is surrounding the point *τ* = 8.17, *η* = 21.86 (Fig.4(e)). The bivariate mixture distribution analysis shows that we can estimate the histogram of *D*_*ik*_ for *i* = 1, 2, …, 14 and *k* = 1, 2, …, 29 using a combination of two bivariate normal distributions, 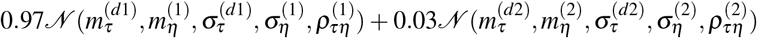 where the superscript 1 (resp. 2) represents the parameters for first (resp. second) component. The parameters of the first component are 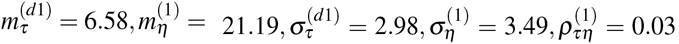, and those of the second component are 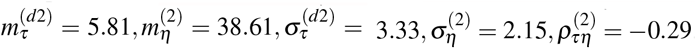.

**Figure 4.**
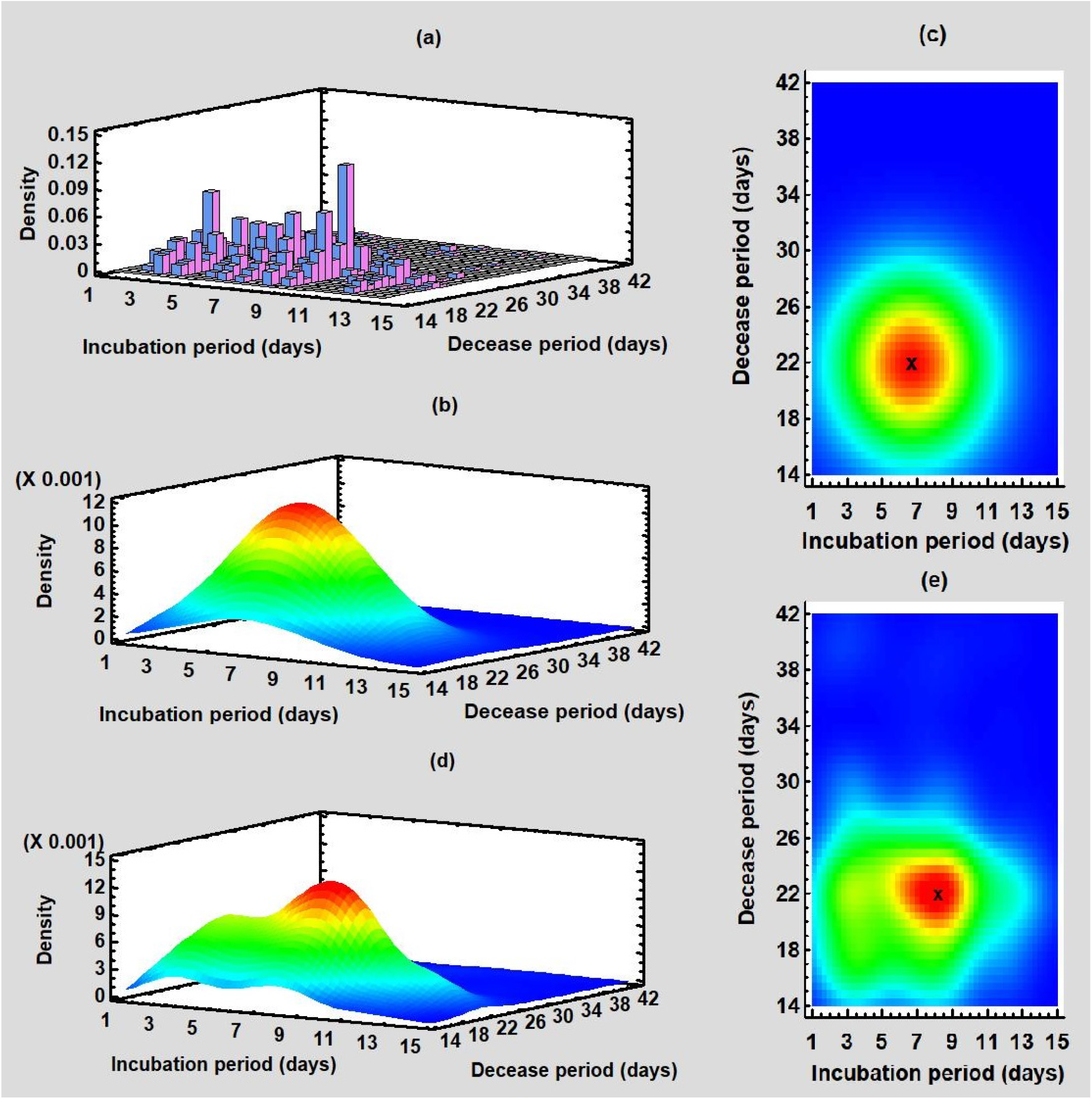
Bivariate distribution of the incubation and decease periods: **(a)** Histogram of the estimated data *D*_*ik*_ for *i* = 1, 2, …, 14 and *k* = 1, 2, …, 29 using the model. **(b)** Fitted bivariate normal distribution. **(c)** Two-dimensional display of (b); the red region is the highly probable domain for decease, and x (6.56, 21.64) denotes the center of the region. **(d)** Fitted nonparametric density estimate with wide 40%; two peaks show that there are two distinguishable high probable regions. **(e)** Two-dimensional display of (d); the x represent the centers of the high probable red region (8.17, 21.86).

### Onset time from symptom to recovery

Using the fact that *τ* + *θ* = *ζ* and the property of expectation *E*(*T* + Θ) = *E*(*T*) + *E*(Θ), we calculate the mean Onset Time from Symptom to Recovery (OTSR) *E*(Θ) (Table1), where *θ* is the variable corresponding to *θ*_*ik*_ for *i* = 1, 2, …, 14 and *k* = 1, 2, …, 29; *T* and Θ are the random variables corresponding the incubation period and OTSR, respectively. There is a good agreement between the calculated values, mean of OTSR, short OTSR and long OTSR, with the reported works (Table1) of earlier studies. However, these calculated values do not show excellent concordance with some other studies, because we consider all recovery cases, mild to moderate, severe, hospitalized (ICU, non-ICU), non hospitalized, in Canada. For example, Voinsky *et. al*.^4^ reported a study with a sample of 5769 patients, not including severe COVID-19 cases. In fact, they mentioned that severe cases were reported to be discharged from the hospital on average 8 days longer than mild to moderate patients requiring hospitalization.

**Table 1.** Comparison of several studies (including the present work) for infectious period along with sample size, mean/median and ranges. Here SD and IQR stand for standard deviation and interquartile range, respectively.

| Author | Location | Data size | Parameter (days) | Reported | Variation (days) | Comment |
| --- | --- | --- | --- | --- | --- | --- |
| Present work | Canada | 72,680 | 15.40 | Mean | 95% CI 14.87 to 15.92 | Onset time from symptom to recovery (OTSR) |
|  |  |  | 14.28 |  | 95% CI 13.79 to 14.77 | Short OTSR |
|  |  |  | 32.14 |  | 95% CI 31.48 to 32.80 | Long OTSR |
| Voinsky *et. al.*<sup>4</sup> | Israeli | 5,769 |  | Mean | 13.24 to 14.81 | Days from first positive to first negative COVID test |
| Barman *et. al.*<sup>5</sup> | India | 221 | 21 | Mean | 95% CI 12.82 to 29.32 | Days of hospitalization Age < 60 yrs. |
|  |  |  | 25 |  | 95% CI 17.22 to 32.78 | Days of hospitalization Age > 60 yrs. |
| Cai *et. al.*<sup>6</sup> | China | 298 | 14 | Median | 9 to 19 (IQR) | Days of treatment within hospital setting |
| Fang *et. al.*<sup>7</sup> | China | 24 | 15.7 | Mean | 6.7(SD) | Days of hospitalization, non-ICU |
| Wu *et. al.*<sup>8</sup> | China | 74 | 16.1 | Mean | 6.7(SD) | Days of hospitalization, severe and non-severe |
| Bi *et. al.*<sup>9</sup> | China | 391 | 21 |  | 95% CI 20 to 22 | Median time to recovery |
| Alinaghi *et. al.*<sup>10</sup> | Iran | 478 | 13.5 |  | IQR: 9 | Median time to recovery |

## Discussion

In the present context, we estimate the recovery as well as decease periods using a novel compartment based model and publicly available database. Here, we consider a maximum length of the incubation period of 14 days, and the ranges of the recovery and decease periods are from 2 to 6 weeks. However, in our method, we can go well beyond all those ranges; the longer ranges simply require a long computational time. Notice that our method could apply the proposed model to estimate key periods for any infectious disease, as along as similar data are available.

The multi-group database *R*_*ik*_, *i* = 1, 2, …, 14 and *k* = 1, 2, …, 29, generated from the model, is the key source to compute all types of distribution of the recovery period, univariate, bimodal and bivariate. The bimodal gamma distribution of the recovery period, 0.9365 Γ(*ζ, K*_*r*1_, *θ*_*r*1_) + 0.0635 Γ(*ζ, K*_*r*2_, *θ*_*r*2_), demonstrates that the recovery period of 93.65% recovered individuals obeys the distribution Γ(*ζ, K*_*r*1_, *θ*_*r*1_), and that of 6.35% recovered individuals obeys the distribution Γ(*ζ, K*_*r*2_, *θ*_*r*2_). Thus, there are two groups of recovered individuals with short recovery period, 21.02 days (on average), and long recovery period, 38.88 days (on average). The characteristics of those two groups may depend on age, underlying health condition, immunity, etc. The database of numerous groups *D*_*ik*_, *i* = 1, 2, …, 14 and *k* = 1, 2, …, 29, generated from the model, is the key source to compute all types of distribution of the decease period, univariate, bimodal and bivariate. The bimodal gamma distribution of the decease period, 0.9508 Γ(*ζ, K*_*d*1_, *θ*_*d*1_) + 0.0492 Γ(*ζ, K*_*d*2_, *θ*_*d*2_), demonstrates that the decease period of 95.08% deaths obeys the distribution Γ(*ζ, K*_*d*1_, *θ*_*d*1_), and that of 4.92% deaths obeys the distribution Γ(*ζ, K*_*d*2_, *θ*_*d*2_). Thus, there are two groups of deaths with short decease period, 21.18 days (on average), and long decease period, 38.41 days (on average). The characteristics of those two groups may depend on age, underlying health condition, immunity, etc. The calculated results employing the proposed model show that the recovery and decease periods are the same. It seems that the survival period of the coronavirus is the same as that of human, in the form of immunity.

The bivariate normal distribution of incubation and recovery periods indicates a recovery window of 4.82 ≤*τ*≤ 8.49 and 19.27 ≤ *ζ* ≤ 25.72 as the highly probable domain for recovery. The bivariate normal distribution of incubation and decease periods indicates a decease window of 4.55 ≤ *τ* ≤ 8.45 and 19.35 ≤ *η* ≤ 24.85 as the highly probable domain of deaths. The study shows that the recovery and decease windows almost coincide within these key periods. To determine precisely the recovery as well as the decease windows, we use nonparametric distributions. Under the nonparametric analysis we identify two recovery windows, 2.27 ≤ *τ* ≤ 4.38, 18.41 ≤ *ζ* ≤ 22.79 and 6.42 ≤ *τ* ≤ 9.63, 17.81 ≤ *ζ* ≤ 23.93, and one decease window, 6.34 ≤ *τ* ≤ 9.41, 20.17 ≤ *η* ≤ 23.69. Nonparametric analysis provides some discrepancy between the recovery and decease windows.

The bivariate mixed distribution, 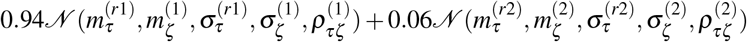, of the incubation and recovery periods demonstrates that 94% recovered individuals obey 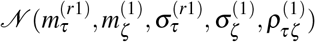, the bivariate normal distribution, with recovery window 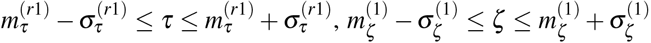 and 6% recovered individuals obey the bivariate normal distribution 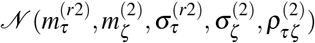 with recovery window 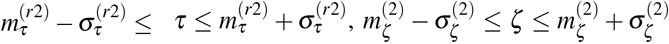. The bivariate mixed distribution, 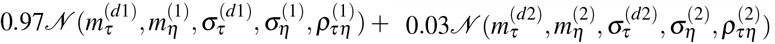, of the incubation and decease periods demonstrates that 97% deaths obey the bivariate normal distribution 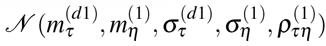 with decease window 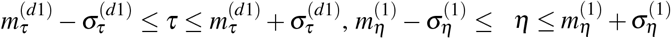 and 3% deaths obey the bivariate normal distribution 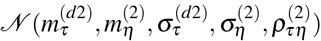 with decease window 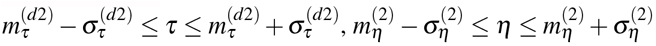.

In summary, we have developed a novel compartment based model to divide the publicly available database of total confirmed cases, recovered cases, and number of deaths into numerous subgroups to obtain univariate and bivariate distributions. The model itself and the procedure to solve it, are the core of this work, and it can be applied to any infectious disease in any region provided that similar data are available. In conclusion, we obtain the distributions of the key periods from the population, considering all types of cases (non-hospitalized, non-ICU, ICU) of recovered individuals and deaths, which is naturally better than any sample-dependent result. In this approach, we do not need any clinical survey; the publicly available data on confirmed cases, recovery and death toll, are sufficient to analyze univariate and bivariate distributions. The current model can be extended to study age-based key periods, and for this purpose we need an age dependent database. The monotonic iteration scheme, introduced for better estimation, can be applied to machine learning as well as numerical analysis problems.

## Methods

In this section, we introduce a compartment based infectious disease model including a large number of partitions, Lockdown, Susceptible, Removed, Infected, fourteen compartments of Confirmed cases, hundreds compartments of Recovered and Deaths. The model is constructed as a set of coupled delay differential equations involving few thousands of variables and parameters, and will be used, not as a prediction tool, but i) for constructing the myriad groups of recovered individuals and death tools and ii) estimating accurately the recovery and decease periods. This model will however have to be parameterized and validated using existing data, in order to justify its accuracy and its application in the proposed methodology.

### The Model

Modeling the spread of epidemic is an essential tool for projecting its outcome. By estimating important epidemiological parameters using the available database and machine learning techniques, we can make predictions of different intervention scenarios. Compartment based model, where the population of a region is distributed into several population groups, such as susceptible, infected, total cases, etc., is a simple but useful tool to demonstrate the panorama of an epidemic.

The proposed model is an extension of our previous work^3^, including a very large number of compartments of recovered and deaths individuals; the schematic diagram of the model is presented in Fig. 5(a). The following are the underlying principles of the present model.

**Figure 5.**
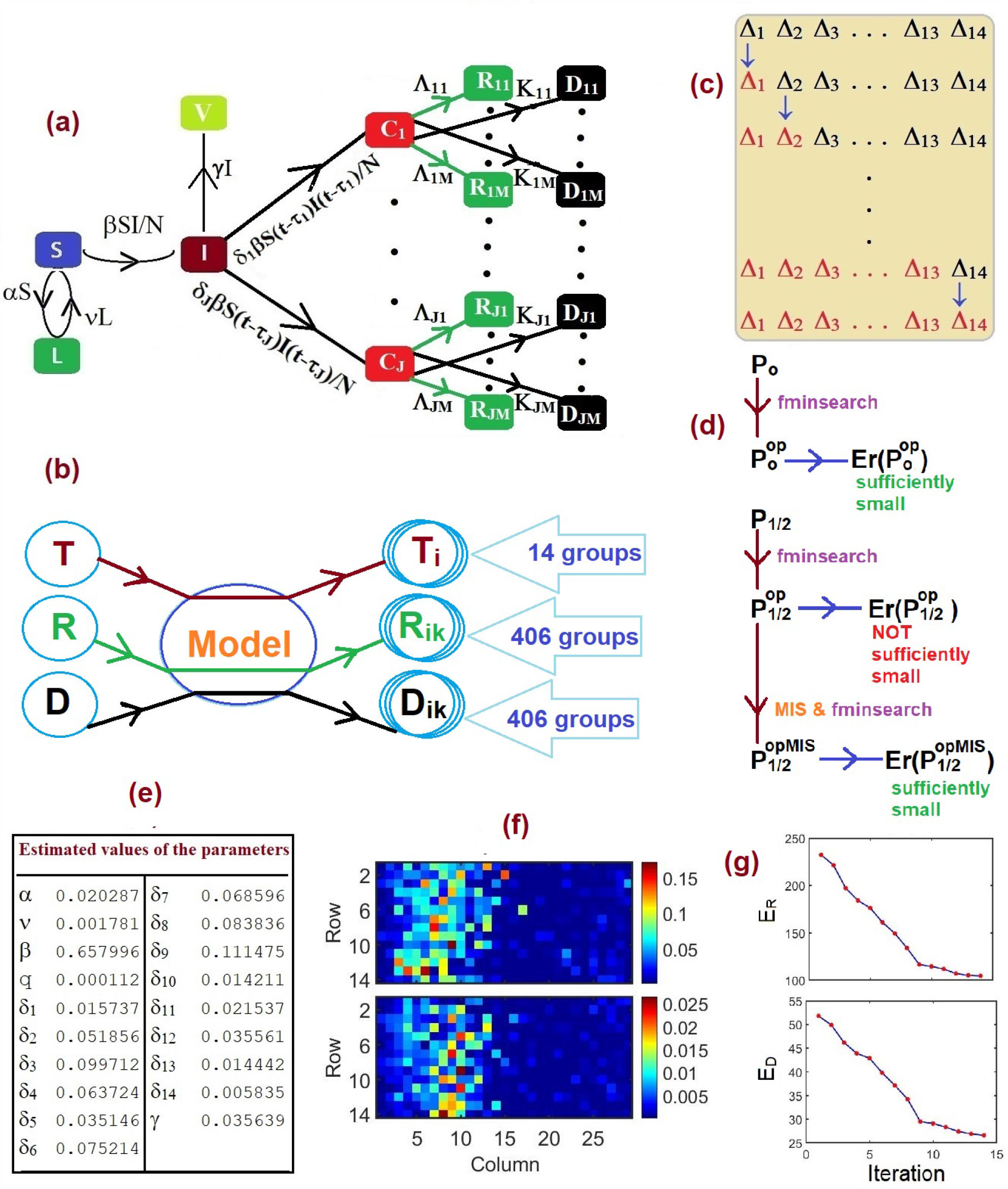
Model, methodology and estimated values of the parameters: **(a)** Schematic diagram of the present compartmental based model, total 830 compartments. Here Λ_*ik*_ = *λ*_*ik*_*δ*_*i*_*βS*(*t* −*τ*_*i*_ −*θ*_*ik*_)*I*(*t* −*τ*_*i*_ −*θ*_*ik*_)*/N* and *K*_*ik*_ = *κ*_*ik*_*δ*_*i*_*βS*(*t* −*τ*_*i*_ −*µ*_*ik*_)*I*(*t* −*τ*_*i*_ −*µ*_*ik*_)*/N* for *i* = 1, 2, …, *J* and *k* = 1, 2, …, *M*. We consider *J* = 14 and *M* = 29. **(b)** Bubble diagram of the foundation of the present work, splitting publicly available database, total cases (T), recovered individuals (R) and death toll (D), into myriad groups. **(c)** Sketch of the Monotonic Iteration Scheme (MIS); for ‘recovery’ calculation Δ_*i*_ ={*λ*_*ik*_| *k* = 1, 2, …, 29} and for ‘decease’ calculation Δ_*i*_ = {*κ*_*ik*_ |*k* = 1, 2, …, 29} and *i* = 1, 2, …, 14. **(d)** Sketch of the optimization scheme for the primary, **P**_0_, and secondary, **P**_1_ and **P**_2_, parameters. **P**_1*/*2_ indicates either **P**_1_ or **P**_2_. **(e)** Estimated values of the primary parameters. **(f)** Estimated values of the secondary parameters, upper panel: *λ*_*ik*_ and lower panel: *κ*_*ik*_. **(g)** Iteration verses error function in MIS, upper panel: estimating *λ*_*ik*_ and lower panel: estimating *κ*_*ik*_.

- The total population is constant (neglecting the migrations, births and unrelated deaths) and initially every individual is assumed susceptible to contract the disease.
- The disease is spread through the direct (face-to-face meeting) or indirect (through air current, common used or delivery items like door handles, grocery products) contact of susceptible individuals with the infected individuals.
- The quarantined area or the compartment for corona cases contains only members of the infected population who are tested corona-positive.
- The virus kills a part of the people it infects; the survivors represent the recovered group.
- There is a non-pharmaceutical policy (stay at home), commonly known as *lockdown*, to stop the spread of the disease.
- The group of asymptomatic patients is a part of infected individuals, and the never-tested recovered asymptomatic patients can be removed from the infected group. If an asymptomatic patient dies, it is counted after investigation.

Based on the above principles, we consider the following compartments:

- Lockdown (insusceptible) (*L*): the group of persons who are keeping themselves safe.
- Susceptible (*S*): the group of individuals who can be infected.
- Infected (*I*): the group of people who are spreading the contiguous disease.
- Removed (*V*): the group of recovered asymptomatic patients without testing.
- Confirmed cases (*C*): the group of individuals who tested corona-positive.
- Recovered (*R*): the group of recovered individuals who tested corona-positive.
- Deaths (*D*): the group of deaths individuals who tested corona-positive.

In the present context, we assume that there is no overlap between these two compartments, infected (*I*) and confirmed cases (*C*). In other words, tested corona-positive individuals are assumed to be unable to substantially spread the disease due to isolation and are immune to re-infection after recovery^30^. The aim of the present work is to estimate the distribution of the recovery and decease periods of COVID-19. In this goal, we split the compartment *C* into *J* subcomponents *C*_1_, …,*C*_*J*_, the compartment *R* into *J ×M* subcomponents *R*_*ik*_ for *i* = 1, …, *J* and *k* = 1, …, *M* and the compartment *D* into *J ×M* subcomponents *D*_*ik*_ for *i* = 1, …, *J* and *k* = 1, …, *M* where

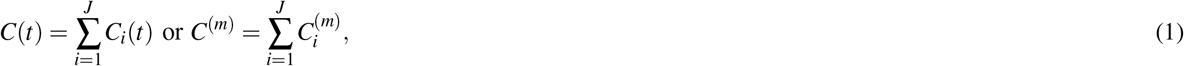

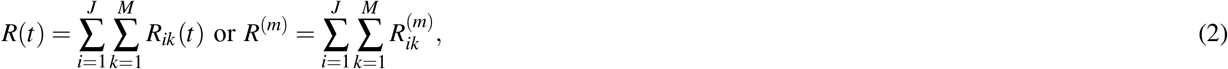

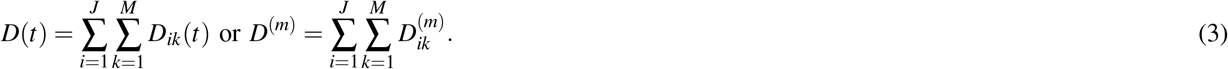

In (1), (2) and (3) *m* represents the time index, and 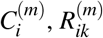 and 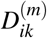 represents the total corona-positive cases corresponding the incubation period *τ*_*i*_, recovered individuals corresponding the incubation period *τ*_*i*_ and onset time *θ*_*ik*_ i.e., recovery period *ζ*_*k*_ = *τ*_*i*_ + *θ*_*ik*_ and death toll corresponding the incubation period *τ*_*i*_ and onset time *µ*_*ik*_ i.e., decease period *η*_*k*_ = *τ*_*i*_ + *µ*_*ik*_, respectively, presented in Fig.5(a).

The time-dependent model is the following set of coupled delay differential equations, for *i* = 1, …, *J*:

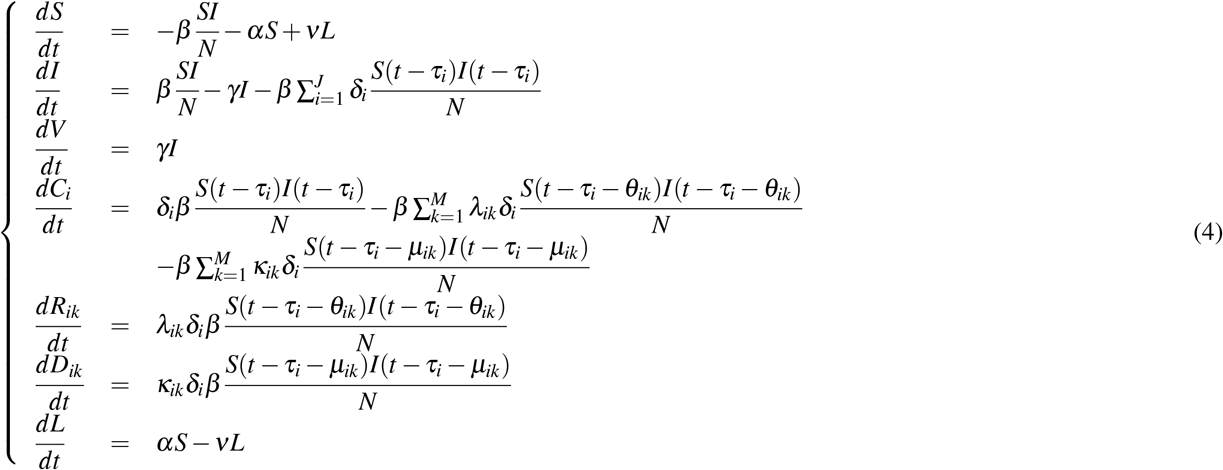

where the real positive modeling parameters *α, β, γ δ*_*i*_, *λ*_*ik*_, *κ*_*ik*_ and *ν* are the rate of lockdown, the rate of infection, the rate of recovery from the asymptomatic group, the rate of tested corona-positive corresponding the incubation period *τ*_*i*_, the rate of recovery corresponding the recovery period *ζ*_*k*_, the rate of decease corresponding the decease period *η*_*k*_ and the rate of transit from lockdown compartment to susceptible compartment, respectively. The variables *S*(*t* −*τ*_*i*_) and *I*(*t* −*τ*_*i*_) denote the cumulative data of (*t* − *τ*_*i*_) days, i.e., total number of suspected and infected individuals of (*t* − *τ*_*i*_) days. The factors *δ*_*i*_*βS*(*t* − *τ*_*i*_)*I*(*t*− *τ*_*i*_)*/N, λ*_*ik*_*δ*_*i*_*βS*(*t* − *τ*_*i*_ − *θ*_*ik*_)*I*(*t* − *τ*_*i*_ − *θ*_*ik*_)*/N, κ*_*ik*_*δ*_*i*_*βS*(*t* − *τ*_*i*_ −*µ*_*ik*_)*I*(*t* −*τ*_*i*_ −*µ*_*ik*_)*/N* convey the rate of individuals who were infected *τ*_*i*_ days ago, the rate of individuals who were infected *τ*_*i*_ + *θ*_*ik*_ days ago and recovered, the rate of individuals who were infected *τ*_*i*_ + *µ*_*ik*_ days ago and died, respectively. It follows from (4), that for any *t*

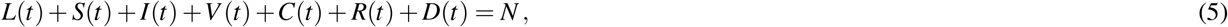

where *N* (constant) is the total population size. We can define a group of new variable *T*_*i*_ for *i* = 1, …, *J* such that

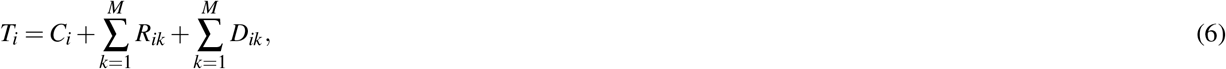

and

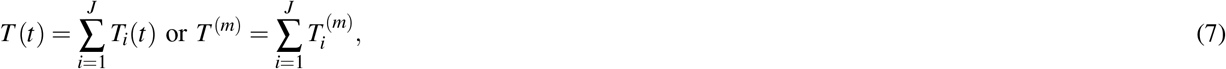

where, *T*, total confirmed cases, is the group of individuals who tested corona positive (active cases + recovered + deaths).

From Eq.(4) we can generate three different sets of coupled delay differential equations for *i* = 1, …, *J* and *k* = 1, …, *M*

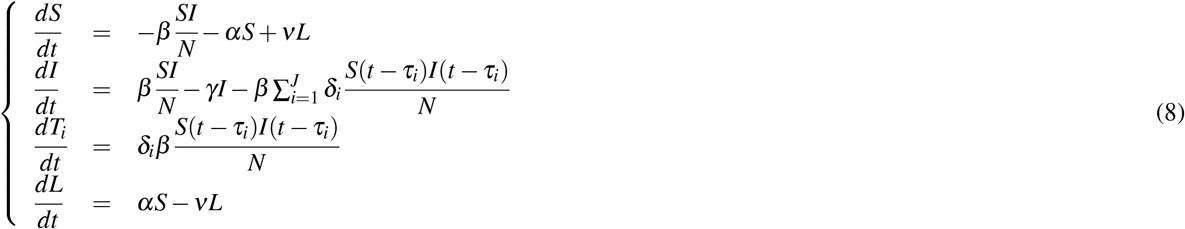

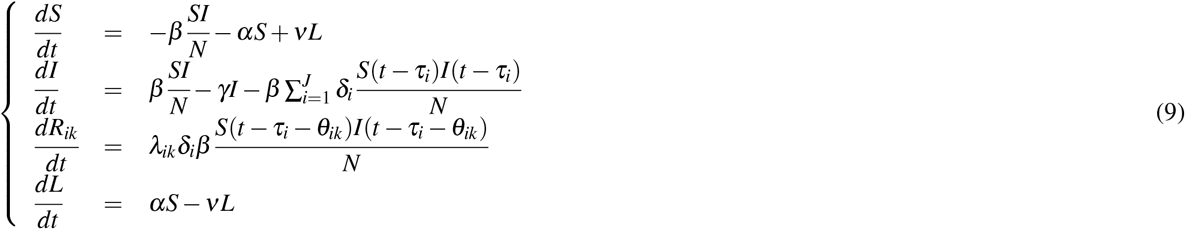

and

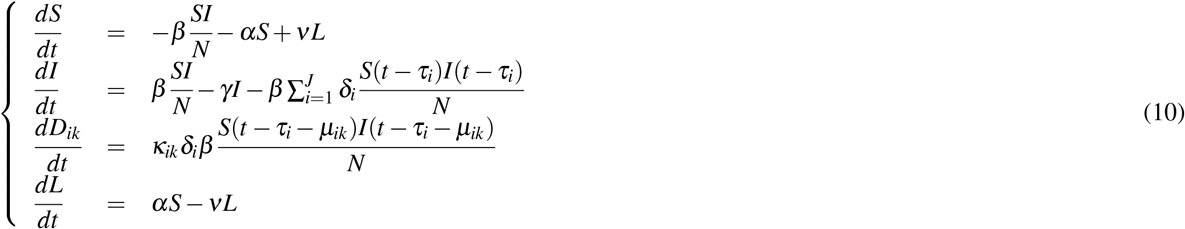

where Eq.(8), (9) and (10) can be used to calculate incubation period^3^, recovery period and decease period, respectively. In the present context, we focus on recovery as well as decease periods. We solve Eq.(8), (9) and (10) using `matlab` inner-embedded function **dde23** with particular sets of model parameters. To solve the initial value problem, in the interval [*t*_0_, *t*_1_], we consider *L*(*t*_0_), *S*(*t*_0_), *I*(*t*_0_), *T* (*t*_0_), *R*(*t*_0_) and *D*(*t*_0_) as follows:

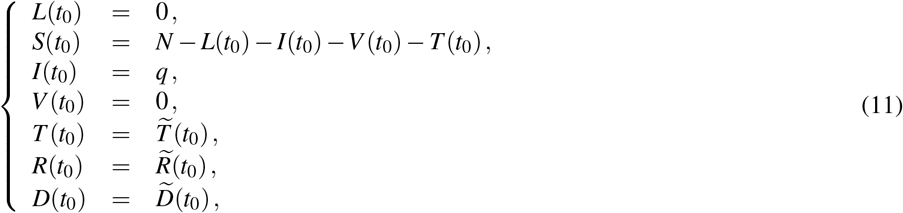

where 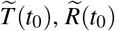 and 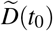 are the available data at time *t*_0_, and *q* is the initial value adjusting parameters. Initially, there are no lockdown individual and no removed individuals from asymptomatic group so that we can consider *L*(*t*_0_) = 0 and *V* (*t*_0_) = 0. It follows from (7) and (11)

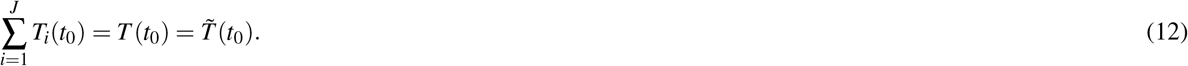

In the present context 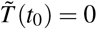, since there were no corona-positive cases reported on January 22, 2020. As a consequence, we also take *T*_*i*_(*t*_0_) = 0 for *i* = 1, 2, …, *J*, and the similar assumptions are valid for *R*_*ik*_(*t*_0_) and *D*_*ik*_(*t*_0_) i.e., *R*_*ik*_(*t*_0_) = 0 and *D*_*ik*_(*t*_0_) = 0 for *i* = 1, 2, …, *J* and *k* = 1, 2, …, *M*.

### Parameter estimation

We focus on the exponential growth phase of the COVID-19 epidemic in Canada; one can use this approach to estimate the incubation, recovery period and decease periods for any region affected by this infectious disease. The time resolved (daily updated) database^27^ provides the number of total corona-positive cases, the number of recovered individuals and the death toll. We define two groups of model parameters: primary parameters, the parameters involved in Eq.(8) i.e., q, *α, β, γ, δ*_*i*_ for *i* = 1, 2, …, *J* and *ν*, and secondary parameters, the parameters involved in Eq.(9) and (10) other than the primary parameters i.e., *λ*_*ik*_ and *κ*_*ik*_ for *i* = 1, 2, …, *J* and *k* = 1, 2, …, *M*. We use the estimated values of the primary parameters to optimize the secondary parameters. The optimal values of the primary parameters **P**_0_ = (*q, α, β, δ*_1_(*t*),, *δ*_*J*_(*t*), *ν*)^*T*^, *q* is the initial value of *I*(*t*), is obtained by minimizing the error function *Er*(**P**_0_), defined as

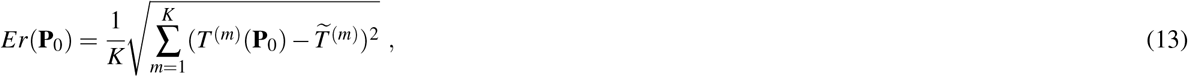

where 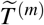 is the available data of total corona-positive cases on the particular *m*th day, and *T*^(*m*)^ is the calculated results obtained from the system (8). The integer *K*, used in (13), is the size of the data set. Due to the complexity of the error function, the minimization using the `matlab` function **fminsearch** requires a very large number of iterations. We use the similar error functions *Er*(**P**_**1**_) and *Er*(**P**_**2**_) to optimize the secondary parameters **P**_**1**_ = (*λ*_11_, …, *λ*_*JM*_)^*T*^ and **P**_**2**_ = (*κ*_11_, …, *κ*_*JM*_)^*T*^, defined as

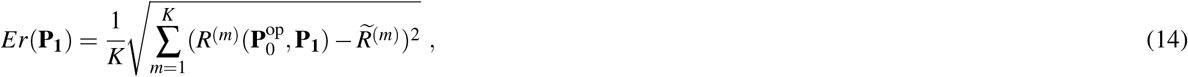

and

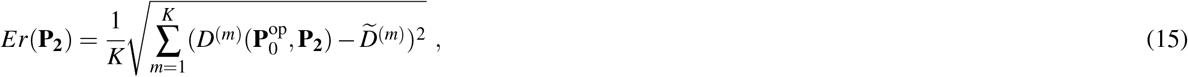

where 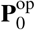 is the estimated values of 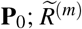 and 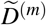 are the available data of total number of recovered individuals and total number of death toll; *R*^(*m*)^ and *D*^(*m*)^ are the calculated results obtained from the Eq.(9) and (10), respectively.

#### Numerical experiment

In this section, we present a detailed description of the computational procedure for the proposed model. On 23 January 2020, a 56-year old man admitted to Toronto hospital emergency department in Toronto with a new onset of fever and nonproductive cough, and returning from Wuhan, China, the day prior^31,32^. It is believed this is the first confirmed case of 2019-nCoV in Canada, and according to the government report, the novel coronavirus arrived on the Canadian coast on January 25, 2020, first reported case. The above information suggests that the start date of the current pandemic in Canada is possibly to be January 22, 2020. Additionally, some research studies reported that the estimation of the incubation period is from 2 days to 14 days, and recovery as well as decease period of COVID-19 is from 2 weeks to 6 weeks^2,33^. As a consequence, in the present study we consider *J* = 14 i.e, 14 delays for the incubation period, and *M* = 29 i.e, 29 delays for the recovery as well as decease periods. Here we consider a calculation of 177 days, from January 22, 2020 to July 16, 2020, duration of the first wave in Canada. The purpose of the model is to separate the publicly available database *T, R* and *D* into myriad groups *T*_*i*_, *R*_*ik*_ and *D*_*ik*_ for *i* = 1, 2, …, 14 and *k* = 1, 2, …, 29 (Fig. 5(b)).

Then the local minimum computed by the optimization algorithm depends on the initial values of the parameters: for *q, α, β, ν* we consider any positive random number less than unity, where as a choice of *δ* = (*δ*_1_, …, *δ*_14_)^*T*^ is tricky. For this purpose, we consider a vector of 14 positive random numbers *δ* such that *δ*_1_ *<* …*< δ*_4_ *< δ*_5_ *> δ*_6_ *>* …*> δ*_14_ and 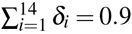. We observe, from numerous numerical experiments, the renormalization factor 0.9 works perfectly for the computation. The estimated values of the primary parameters 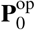 are presented in Fig.5(e), and the value of the error function *Er*(**P**_0_) = 41.64. The estimated values of the primary parameters are related to Eq.(8), the set of coupled delay differential equations, and Eq.(13), the error function. Using the estimated values of the primary parameters, we optimize the secondary parameters *λ*_*ik*_ for *i* = 1, 2, …, 14 and *k* = 1, 2, …, 29 related to Eq.(9) and (14). The choice of the initial values of *λ*_*ik*_ is such that for any fixed *i, i* = 1, 2, …, 14, the first fourteen *λ*_*ik*_s i.e., *{λ*_*i*1_, *λ*_*i*2_, …, *λ*_*i*14_*}* are in ascending order, and the rest i.e., *{λ*_*i*15_, *λ*_*i*16_, …, *λ*_*i*29_*}* are in descending order; and 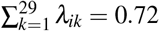. After optimization, we obtain the value of the error function *Er*(**P**_**1**_^op^) = 236.47. Using the estimated values of the primary parameters, we optimize the secondary parameters *κ*_*ik*_ for *i* = 1, 2, …, 14 and *k* = 1, 2, …, 29 related to Eq.(10) and (15). The choice of the initial values of *κ*_*ik*_ is such that for any fixed *i, i* = 1, 2, …, 14, the first fourteen *κ*_*ik*_s i.e., *{κ*_*i*1_, *κ*_*i*2_, …, *κ*_*i*14_*}* are in ascending order, and the rest i.e., *{κ*_*i*15_, *κ*_*i*16_, …, *κ*_*i*29_*}* are in descending order; and 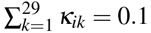. After optimization, we obtain the value of the error function 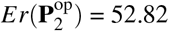. The values of the error functions 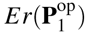 and 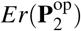 are not sufficiently small. To overcome that difficulties, here, we introduce a Monotonic Iteration Scheme (MIS).

#### Monotonic Iteration Scheme

To optimize the parameters *ξ*_*ik*_ for *i* = 1, 2, …, *J* and *k* = 1, 2, …, *M*, we use a MIS with *J* = 14 and *M* = 29. However, the method can be applied for any finite integer values of *J* and *M*. The schematic diagram of MIS is presented in Fig.5(c), and consists of the following steps.

- **Step 1:** We decompose the parametric domain Δ = *{ξ*_*ik*_|*i* = 1, 2, …, 14; *k* = 1, 2, …, 29*}* into 14 subdomains Δ_*i*_ = *{ξ*_*ik*_|*k* = 1, 2, …, 29*}* so Δ = *{*Δ_1_, Δ_2_, Δ_3_, …, Δ_14_*}*.
- **Step 2:** We optimize the subdomain Δ_1_ and consider the other parameters Δ_2_, Δ_3_, …, Δ_14_, as constants. After first iteration, we get estimated parameters 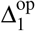; the entire parametric domain is 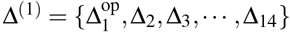, and the error function *Er*(Δ^(1)^).
- **Step 3:** In the second iteration, we optimize the subdomain Δ_2_ and keeping the other subdomains of Δ^(1)^ unchanged. After second iteration, we get estimated parameters 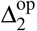; the entire parametric domain is 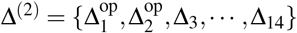, and the error function *Er*(Δ^(2)^).
- **Step 4:** Repeated the same procedure discussed in **Step 3**.

The optimization of the subdomain Δ_2_, demonstrated in **Step 3**, is related to minimizing the error function such that *Er*(Δ^(1)^) *≥ Er*(Δ^(2)^); the equality sign holds for 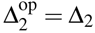. The error function of the *n* + 1th iteration, *Er*(Δ^(*n*+1)^) cannot be greater than that of *n*th iteration, *Er*(Δ^(*n*)^), because of this characteristic of the error function we define the approach as MIS. The flow chat of the optimization scheme is presented in Fig. 5(d). The upper and lower panels of Fig.5(f) show the estimated values of the secondary parameters *λ*_*ik*_ and *κ*_*ik*_, respectively, obtained from the MIS. The upper and lower panels of Fig.5(g) show the values of the error functions *E*_*R*_, using MIS to optimize *λ*_*ik*_, and *E*_*D*_, using MIS to optimize *κ*_*ik*_, for fourteen iteration steps and 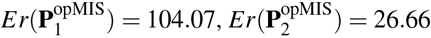 where 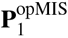 and 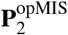 are the estimated values of 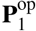 and 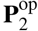, respectively, using MIS. Fig. 5(g) shows that the MIS works efficiently to get better estimations.

## Data Availability

The data used to estimate the model parameters are publicly available and are available with the code.

https://resources-covid19canada.hub.arcgis.com/datasets/case-accumulation/data

## Code availability

All code is available in the GitHub repository for the project at https://github.com/SPAUL2021/COVID19RECOVERY/tree/SPAUL2021-patch-1

## Acknowledgements

This research is supported by the Natural Sciences and Engineering Research Council of Canada (NSERC).

## Author contributions statement

S.P. has derived the model, has developed the matlab code, has analyzed the calculated results, and has prepared all figures and tables. S.P. and E.L. have drafted the original article. Both authors have contributed to the editing of the article. Both authors have read and approved the final article.

## Additional information

Correspondence and requests for materials should be addressed to S.P.

